# Innovations to improve access to outpatient rehabilitation services for persons presenting physical disabilities: results of an environmental scan in Quebec, Canada

**DOI:** 10.1101/2025.09.08.25335182

**Authors:** Geneviève Ferland, Frédérique Dupuis, Nathan Blanchard, Élizabeth Marcoux, Marie-Ève Lamontagne, Anne Hudon, Luc J. Hébert, Jean-Sébastien Roy, Angel Ruiz, Anne Marie Pinard, François Routhier, François Desmeules, Kadija Perreault

## Abstract

**PURPOSE:** To identify and characterize innovations implemented to improve access to publicly funded outpatient rehabilitation services for people experiencing physical disabilities, and to explore main barriers and facilitators to implementation.

**MATERIALS AND METHODS:** An environmental scan was conducted in the province of Quebec (Canada), combining an online search and semi-structured interviews with key informants. For each innovation, data on implementation context, targeted access issue(s), and indicators used to measure impact were extracted and analyzed descriptively. Interviews were transcribed and analyzed using a mixed inductive/deductive thematic approach to identify barriers and facilitators.

**RESULTS:** Twenty-seven innovations, implemented in various regions and settings, were identified. Most of these targeted individuals with musculoskeletal disorders and addressed issues such as long wait times, geographical inequalities, and inappropriate care. Interviews were conducted with informants (n=26) involved in 20 of the innovations. Main barriers to implementation included divergences between actors’ perspectives on the relevance of innovations, their complexity and insufficient resources. Main facilitators were having strong planification and individual’s engagement in the implementation.

**CONCLUSIONS:** This study enhances our understanding of the nature and purpose of innovations aimed at improving access to outpatient rehabilitation for people with physical disabilities. Implementation is a complex process that requires careful consideration of multiple factors.

**Implications for Rehabilitation:**

- The results highlight multiple innovations that have been implemented to improve access to rehabilitation services, which may help inform settings seeking such innovations.
- Identified innovations targeted various dimensions of access, including wait times, geographic barriers, service appropriateness, and equity.
- Implementing such innovations should consider numerous factors, including administrative burden and the need for adequate technical support.

## INTRODUCTION

Difficulties in accessing publicly funded healthcare services represent a major preoccupation in many countries. Recurrent manifestations of limited access include long waiting times and disparities in access between subgroups of populations (1–6). Access to healthcare results from the interface between the 1) *abilities of* individuals, their households and communities to perceive, seek, reach, pay and engage in healthcare, and the 2) *characteristics* of services, providers and systems (7).

Lack of access to healthcare has mostly been documented in medical (e.g. primary care, emergency departments) and hospital-based services (4,6,8,9). In contrast, evidence in rehabilitation remains sparse (10,11), although physical disabilities are the leading type of invalidity worldwide (12). Important limitations in access to rehabilitation have been identified, such as waiting times (1,13,14). Notably, long waiting times to rehabilitation services have been associated with deterioration of health, quality of life and psychological well-being (2,15–17). In 2019, the World Health Organization launched the Rehabilitation 2030 initiative in order to focus attention on the “*profound unmet need for rehabilitation in the world*” (18), emphasizing that “*rehabilitation is a necessary health service that should be part of universal health coverage, at all levels of healthcare, and available for all populations and through all stages of the life course*”(12).

Hence, identifying strategies to improve access to rehabilitation services is a crucial issue that merits attention in any healthcare systems. A systematic review conducted by Dupuis and al. assessed the effectiveness of service redesign strategies to reduce waiting times before accessing outpatient rehabilitation services for adults with physical disabilities (19). Based on the findings of this review, effective strategies to reduce waiting times fall under three categories: strategies that target improvements in access processes and referral management, those that introduce new or modified models of care delivery and those that extend or modify the roles and scope of practice of allied-health professionals. It is likely that other relevant innovations or strategies have been developed within healthcare systems without being systematically documented, published or investigated using a scientific approach. Identifying such innovations is a way to learn from past experiences, prevent reinventing the wheel, and guide decision-making to help improve access to rehabilitation.

To our knowledge, no previous work has been carried out to document innovations that have already been implemented to improve access to rehabilitation services in the province of Quebec, Canada’s second most populated province. Therefore, the first objective of this study was to identify and characterize innovations implemented to improve access to publicly funded outpatient rehabilitation services for persons experiencing physical disabilities. The second objective was to explore the main barriers and facilitators to the implementation of these innovations.

## MATERIALS AND METHODS

### Study design

This project used an environmental scan design combining a structured online search with semi-structured interviews with key informants. Environmental scans combine diverse sources of data (e.g. website, report) and have been used increasingly for diverse purposes, including health services decision-making (20,21). For instance, they are used to inform about current trends and contexts of practice, and to plan the development of programs or policies that align with the health needs of communities (20). Since this project used publicly available data as well as interviews with key informants that focused on implemented innovations, ethics approval was considered unnecessary by the institutional ethics committee.

### Data collection

#### Online search

Innovations were first identified through an online search. Our search process first involved the preparation of a list of online sources that could potentially serve to identify innovations implemented in Quebec. This list included websites of grey literature, including Quebec healthcare organizations and websites specifically reporting innovations in healthcare, and the Google search engine (21–24). The online research was performed between June 2021 and November 2022, and included documents published after 2015, year of a major reform in Quebec’s health system focusing on the reorganization of health and social service institutions (25,26). All search queries were performed using an «in-cognito» page to avoid personalization bias (23). The keywords used to identify the innovations were related to three main concepts: access, innovations and type of rehabilitation service (e.g. occupational therapy, physiotherapy, speech therapy, etc.) (see Appendix 1). For each search query, the first 100 documents were screened for eligibility. Documents published in English or French were considered. In addition to these online searches, innovations identified through subsequent interviews were also considered for inclusion.

Documents initially identified through the online search were then screened to verify if the innovation answered these additional criteria: 1) it was coherent with the definition of innovation as *an intervention that changes the organization and/or delivery of services at any point in the patient journey* (27); 2) it aimed to improve access to rehabilitation services; 3) it was implemented in publicly funded outpatient care settings in the province of Quebec; and 4) it targeted services for adults presenting physical disabilities. Each innovation was screened for eligibility by two independent reviewers (GF and FDu). Discrepancies between reviewers were resolved by consensus or else a third reviewer was consulted (KP).

For each retained innovation, data were manually extracted from the documents and synthesized in a data extraction grid. To answer the first aim of the study, a description of the context in which the innovations were implemented was performed. The context included 1) the location of the service, 2) the targeted population, 3) the clinical setting, and 4) the rehabilitation professionals involved. A description of the nature of the innovations was also performed, which included 1) the type of innovation according to Dupuis et al.’s categorization (19), 2) the targeted access-related problem, according to the five dimensions of accessibility described by Lévesque et al.(7) (i.e., approachability, acceptability, availability and accommodation, affordability, and appropriateness), and 3) the scale of the innovations (provincial, organizational, facility, or service-level). In addition, a description of the indicators used to document any changes on access were extracted, when available.

#### Semi-structured interviews with key informants

Following the identification of innovations, semi-structured interviews were conducted with key informants that participated in the implementation of the innovations included. The interviews took place between August and November 2022 and were carried out by one or two team members (GF/KP). This step was meant to allow a more detailed and comprehensive understanding of the innovations, by filling the gaps in information obtained from the other data sources. Barriers and facilitators to the implementation of innovations were also addressed.

Key informants were identified through contact details in available documents or, when unavailable, by reaching out to the organizations responsible for the innovation to locate individuals directly involved in its implementation. When contact with key informants was successfully achieved (by phone or email) and their consent was obtained, a conference call using the Zoom platform (Zoom Video Communications, San Jose, CA, USA) was planned. An interview guide was designed beforehand and covered: underlying obstacles to access and motives for innovation development, core characteristics of implemented innovations, and barriers and facilitators to implementation. The domains and subdomains of the Consolidated Framework for Implementation Research (CFIR) were used to inform the questions regarding barriers and facilitators (28). Key informants were also asked to identify any other implemented innovations they were aware of, to include any relevant missing innovations. The interviews lasted between 45 and 75 minutes, were recorded, and their content was transcribed verbatim by a professional firm.

### Data analysis

Data extracted through the documentary analysis was synthesized using descriptive statistics. The transcriptions of the interviews with key informants were first verified for exactness (29). Additional data obtained through the interviews regarding the innovations themselves were included in the data extraction grid. The content of the interviews was then analyzed using a mixed inductive and deductive qualitative thematic analysis (30) to identify main barriers and facilitators. Transcriptions were initially coded deductively using the domains and subdomains of the CFIR (28). Then a grid was designed to help visualize key content grouped according to the domains and subdomains. We then carried out a more inductive level of analysis, by identifying patterns through the grid, which led to the identification of main themes. The themes were first developed by one author (GF) and refined based on discussions and analysis sessions with team members. The NVivo 12 software was used to conduct the qualitative analysis (31).

## RESULTS

### Search results

The online search originally identified 158 innovations. After reading full documents, 31 innovations were included. The last step of validation of the selection criteria (i.e., the innovation targeted services for adults presenting physical disabilities) was carried out when contacting key stakeholders, resulting in 27 innovations included (Figure 1). Different types of data sources were identified on the websites, including internal newsletters (n=10), research project reports (n=8), publicly available service descriptions (n=6), newspaper articles (n=3), promotional videos (n=2), conference/presentations (n=2), and annual reports (n=1). The semi-structured interviews were conducted with 26 key informants for 20 of the included innovations, with six interviews that were conducted with two key informants at the same time. No additional innovations were identified through the interviews. An overview of innovations is presented in Table 1, while Table 2 provides characteristics of each retained innovation.

**Figure 1.**
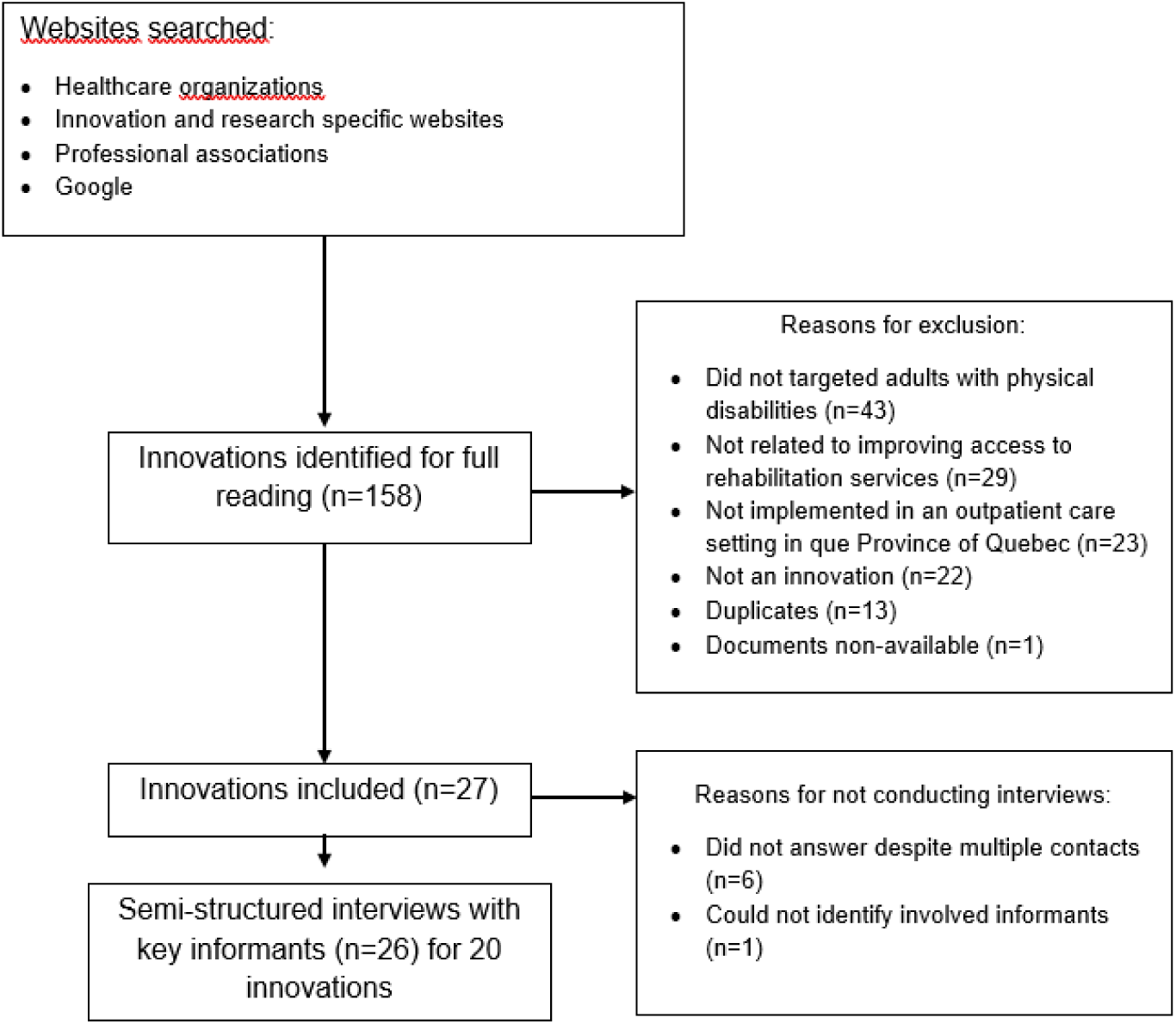
Flow chart of online search and identification of innovations

**Table 1.**
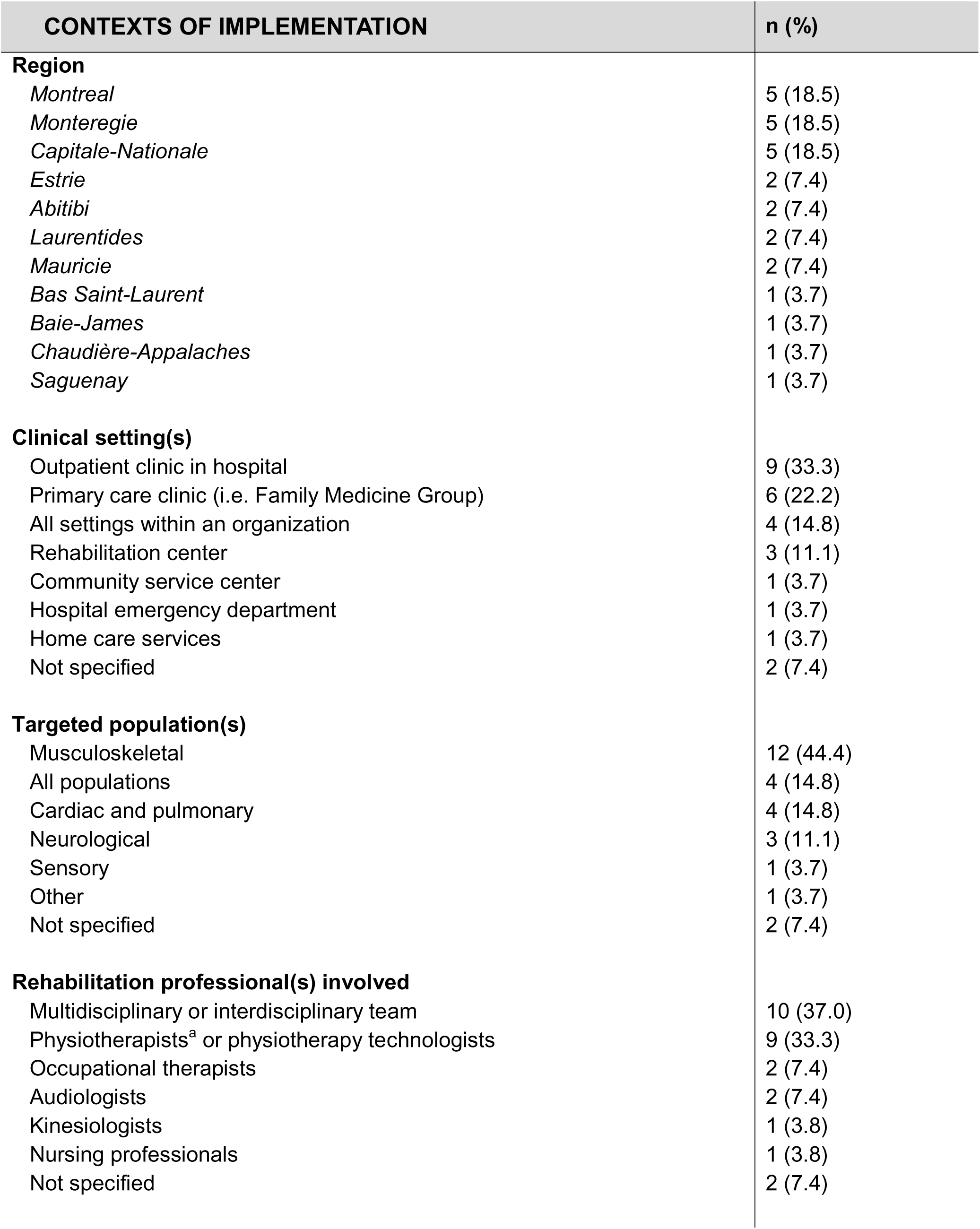
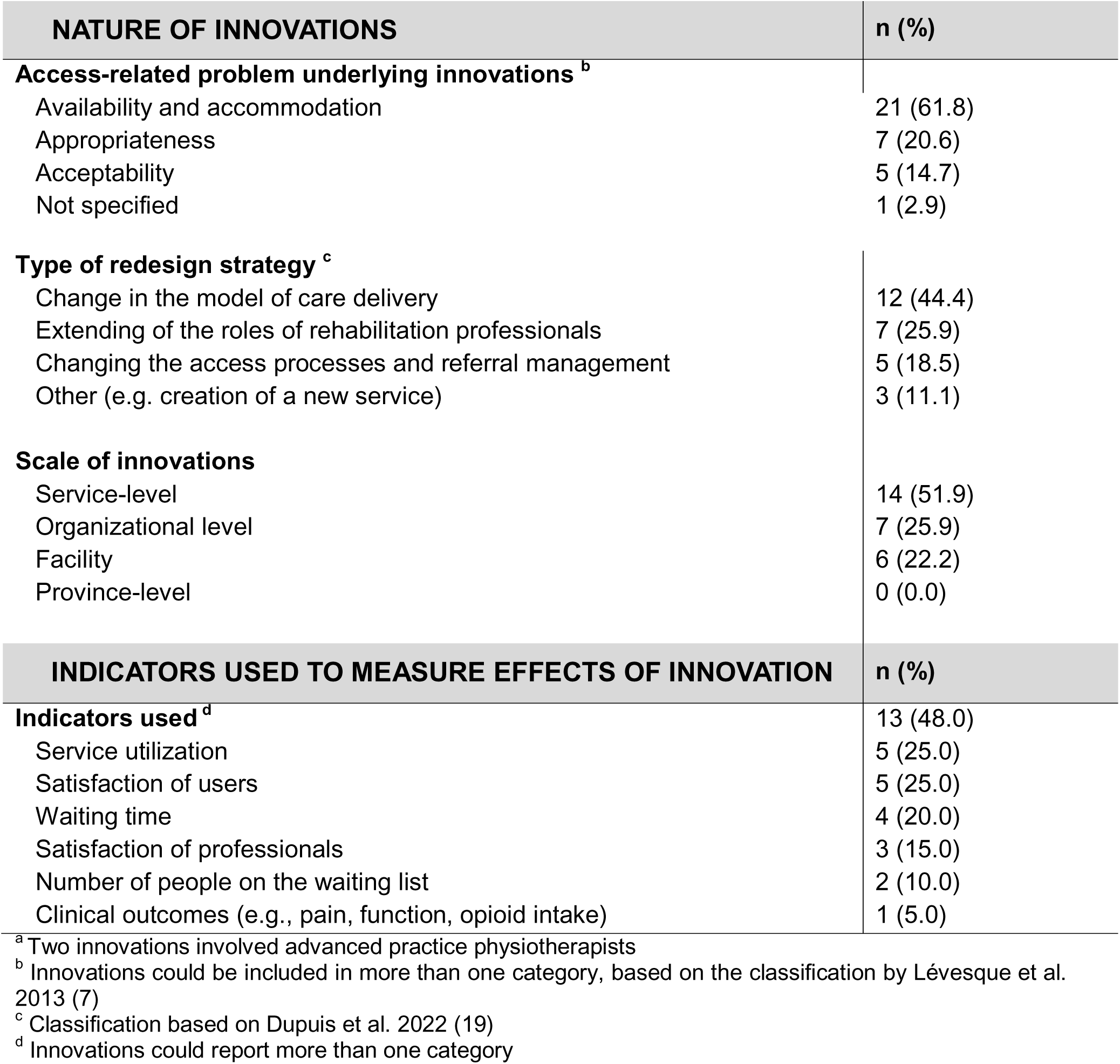
Overview of innovations identified based on their characteristics (n=27 unless mentioned otherwise)]

| CONTEXTS OF IMPLEMENTATION | n (%) |
| --- | --- |
| **Region** |  |
| *Montreal* | 5 (18.5) |
| *Monteregie* | 5 (18.5) |
| *Capitale-Nationale* | 5 (18.5) |
| *Estrie* | 2 (7.4) |
| *Abitibi* | 2 (7.4) |
| *Laurentides* | 2 (7.4) |
| *Mauricie* | 2 (7.4) |
| *Bas Saint-Laurent* | 1 (3.7) |
| *Baie-James* | 1 (3.7) |
| *Chaudière-Appalaches* | 1 (3.7) |
| *Saguenay* | 1 (3.7) |
| **Clinical setting(s)** |  |
| Outpatient clinic in hospital | 9 (33.3) |
| Primary care clinic (i.e. Family Medicine Group) | 6 (22.2) |
| All settings within an organization | 4 (14.8) |
| Rehabilitation center | 3 (11.1) |
| Community service center | 1 (3.7) |
| Hospital emergency department | 1 (3.7) |
| Home care services | 1 (3.7) |
| Not specified | 2 (7.4) |
| **Targeted population(s)** |  |
| Musculoskeletal | 12 (44.4) |
| All populations | 4 (14.8) |
| Cardiac and pulmonary | 4 (14.8) |
| Neurological | 3 (11.1) |
| Sensory | 1 (3.7) |
| Other | 1 (3.7) |
| Not specified | 2 (7.4) |
| **Rehabilitation professional(s) involved** |  |
| Multidisciplinary or interdisciplinary team | 10 (37.0) |
| Physiotherapists<sup>a</sup> or physiotherapy technologists | 9 (33.3) |
| Occupational therapists | 2 (7.4) |
| Audiologists | 2 (7.4) |
| Kinesiologists | 1 (3.8) |
| Nursing professionals | 1 (3.8) |
| Not specified | 2 (7.4) |

| NATURE OF INNOVATIONS | n (%) |
| --- | --- |
| **Access-related problem underlying innovations <sup>b</sup>** |  |
| Availability and accommodation | 21 (61.8) |
| Appropriateness | 7 (20.6) |
| Acceptability | 5 (14.7) |
| Not specified | 1 (2.9) |
| **Type of redesign strategy <sup>c</sup>** |  |
| Change in the model of care delivery | 12 (44.4) |
| Extending of the roles of rehabilitation professionals | 7 (25.9) |
| Changing the access processes and referral management | 5 (18.5) |
| Other (e.g. creation of a new service) | 3 (11.1) |
| **Scale of innovations** |  |
| Service-level | 14 (51.9) |
| Organizational level | 7 (25.9) |
| Facility | 6 (22.2) |
| Province-level | 0 (0.0) |
| INDICATORS USED TO MEASURE EFFECTS OF INNOVATION | n (%) |
| **Indicators used <sup>d</sup>** | 13 (48.0) |
| Service utilization | 5 (25.0) |
| Satisfaction of users | 5 (25.0) |
| Waiting time | 4 (20.0) |
| Satisfaction of professionals | 3 (15.0) |
| Number of people on the waiting list | 2 (10.0) |
| Clinical outcomes (e.g., pain, function, opioid intake) | 1 (5.0) |

<sup>a</sup> Two innovations involved advanced practice physiotherapists
<sup>b</sup> Innovations could be included in more than one category, based on the classification by Lévesque et al. 2013 (7)
<sup>c</sup> Classification based on Dupuis et al. 2022 (19)
<sup>d</sup> Innovations could report more than one category

**Table 2.**
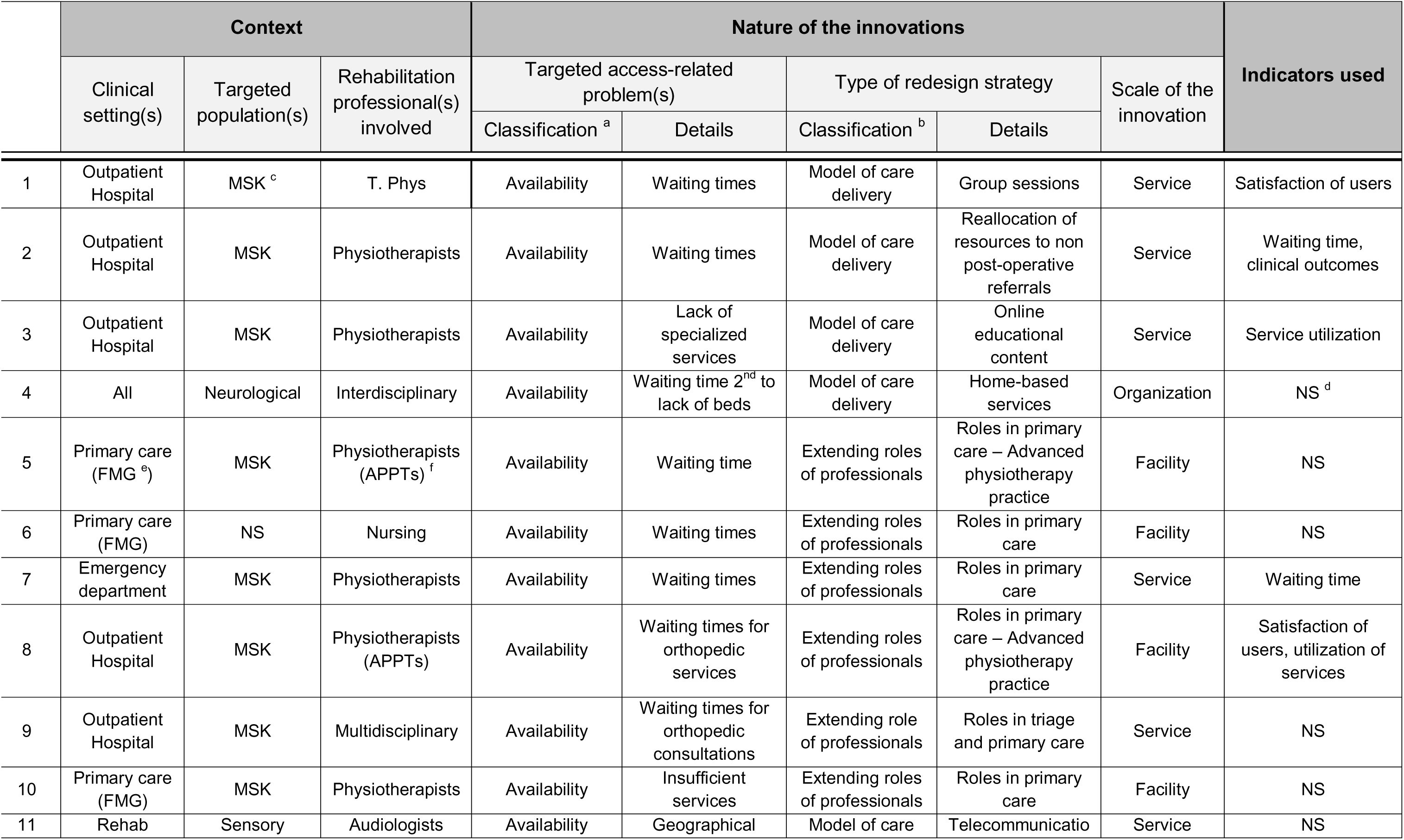
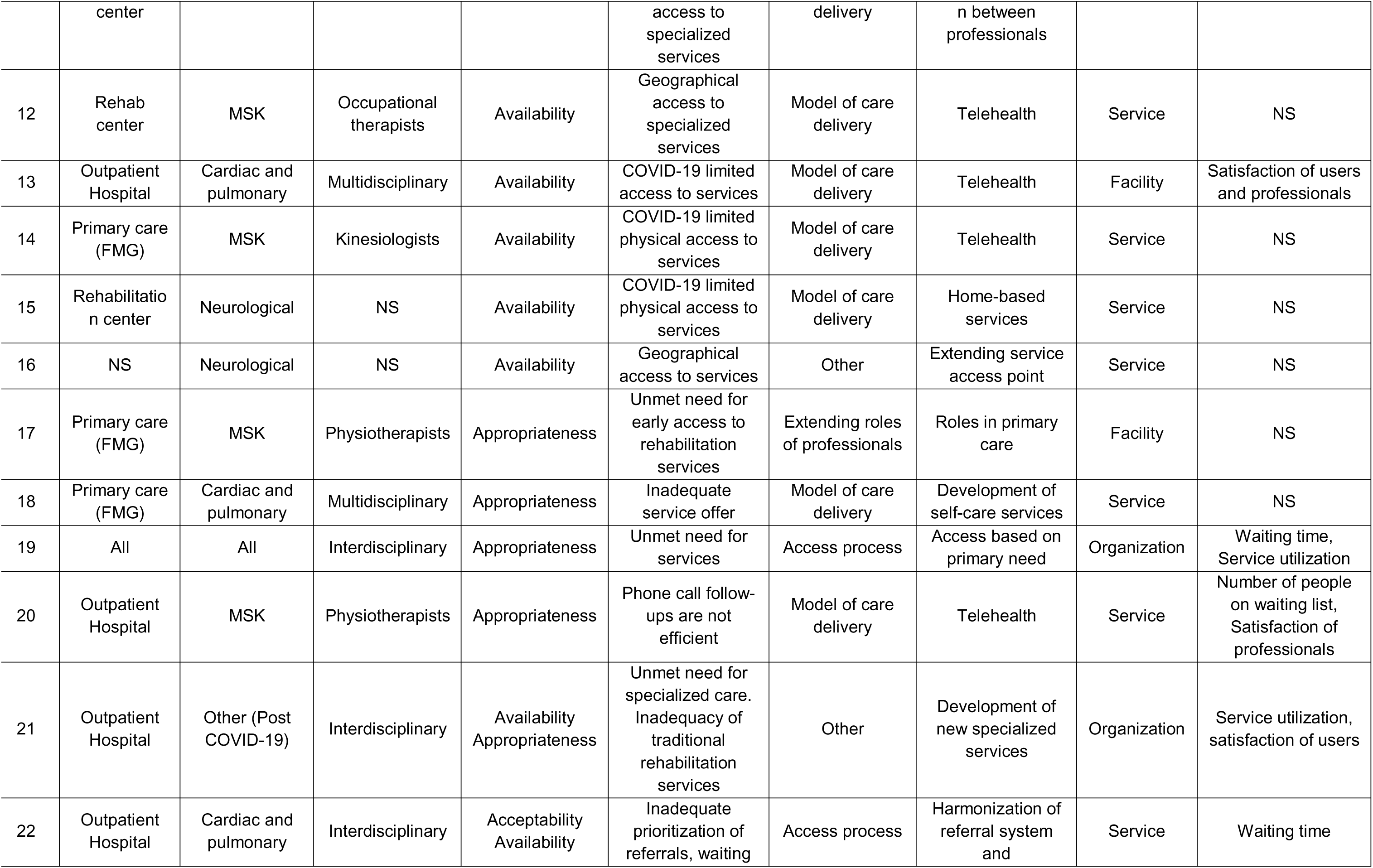
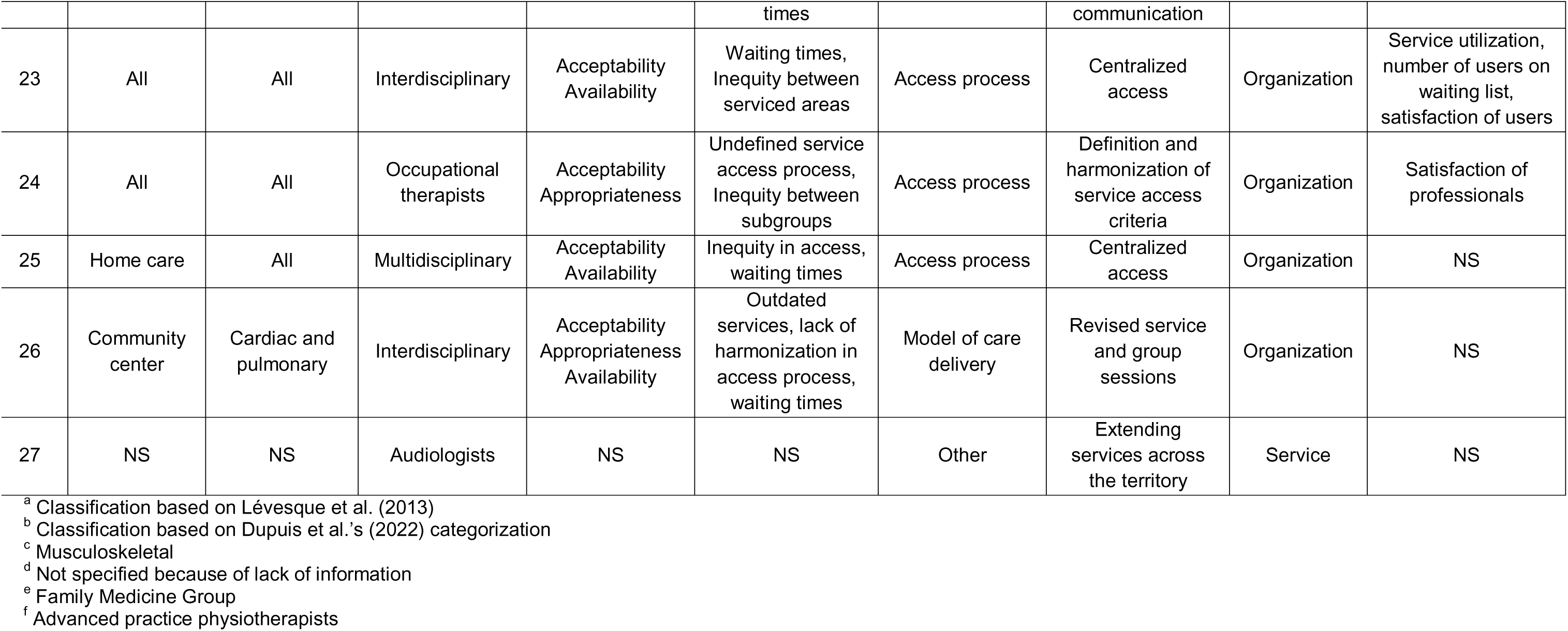
Detailed description of included innovations (n=27)

### Contexts of implementation

Most innovations were observed in three large and urban health regions: *Montréal* (19%)*, Montérégie* (19%) and *Capitale-Nationale* (19%). People presenting musculoskeletal disorders were the most targeted population with 48% of all included innovations. Five innovations (15%) did not target a specific population; they aimed to serve all adults with physical disabilities. There was great disparity in the type of healthcare settings where the innovations were implemented. Primary care clinics (e.g. *Family Medicine Groups*) and outpatient clinics were the most frequent. There was often more than one type of rehabilitation professional involved in the targeted services (37%), either in multidisciplinary or interdisciplinary contexts. When only one professional was involved, physiotherapists or physiotherapy technologists were the most represented (33%). The detailed characteristics of the contexts of implementation are reported in Table 1.

### Nature of innovations

Based on Lévesque et al.’s (7) access to health care framework*, availability and accommodation* were the main access-related issues that were targeted through implementation (62%) of the innovations. Specifically, unacceptable waiting times limiting access to services were most frequently reported. Based on Dupuis et al.’s categorization, 44% of the innovations were classified as bringing a change in the model of care delivery (19). Among these, a considerable number of innovations modified the location of service delivery, mainly introducing telerehabilitation. Extending or modifying the roles of rehabilitation professionals were at the heart of 26% of innovations, mainly through the introduction of screening and care led by rehabilitation professionals in settings that were traditionally medical, such as Family Medicine Groups, emergency departments or orthopedic clinics. In two innovations, extending roles of professionals translated into the introduction of advanced-practice (APP) models of care. Changes in access processes and referral management, such as harmonization and centralization of access, was a strategy proposed in 19% of innovations (Table 1). Other innovations (11%) could not be categorized into these three categories; they involved the implementation of new services, i.e. the addition of a new service to provide care for a specific patient population, and the expansion of a rehabilitation service in underserved areas. Innovations were most frequently implemented at the service level (52%; e.g. physiotherapy group session for post-operative patients in an outpatient external clinic).

### Use of indicators measuring the effects of the innovations

For nearly half of the innovations (48%), data was collected to evaluate the effects of the innovation. A variety of indicators were used, the most frequent being service utilization (i.e. number of patients receiving the service) (25%), satisfaction surveys conducted with users (25%) and waiting times (20%).

### Barriers and facilitators to the implementation of the innovations

Several barriers and facilitators to the implementation of the innovations were identified through the semi-structured interviews. The barriers fell under three main themes: 1) divergence between actors’ perspectives on the relevance of the innovation, 2) operational complexity of care model transformation, and 3) insufficient resources to achieve successful implementation. The facilitators were classified under two main themes: 1) matching evidence, contextual needs and resources, and 2) the quality of the team involved in leading and conducting the implementation process.

#### Barrier 1 - Divergence between actors’ perspectives on the relevance of the innovation

Discrepant perspectives among different stakeholder groups (e.g. type of professionals, between managers and clinicians) represented a major barrier, particularly when the added value of the innovation was unclear or when concerns about its safety and impact on clinical practice were raised. Such divergence was said to create some resistance when implementing innovations, especially for innovations that modified or extended the roles of rehabilitation professionals. For instance, innovations where physiotherapists were involved in primary care delivery, such as the deployment of direct-access physiotherapy within emergency department and Family Medicine Groups or through APP models of care. Those were generally associated with disbeliefs, fears and doubts towards the competency of rehabilitation professionals, thereby raising concerns about the acceptability of such innovations. As one informant mentioned:

> *“The medical community might agree, but they still question whether physiotherapists are truly capable of managing these patients. They worry about potential diagnostic errors or the possibility of missing something serious. Therefore, this concern about physiotherapists’ competence originates from the medical teams.” (Key informant 13)*

Certain key informants also discussed that having previous positive experiences with the extension or modification of rehabilitation professionals’ roles could potentially diminish people’s concerns and resistance.

Diverging perspectives were reported between rehabilitation professionals—who believed they should have greater clinical autonomy in primary care—and other clinicians, particularly physicians, as well as other key stakeholders, who expressed resistance to expanding their role and scope practice. According to several informants, this lack of consensus limited the recognition and integration of rehabilitation professionals in new care models, thereby slowing down implementation efforts and undermining the potential impact of these innovations on access improvement:

> *“I noticed that this deviation was occurring (…) given that we don’t have professional autonomy to, for instance, prescribe an X-ray, complete a workers’ compensation form, fill out insurance documents, or work-related sick leave — these limitations significantly restricted our ability to intervene. (…)”* (Key informant 20).

#### Barrier 2 - Operational complexity of care model transformation

Another barrier concerned the operational complexity involved in implementing and integrating these innovations into the daily practice of rehabilitation professionals. For example, modifying the model of care delivery sometimes involved the use of new technologies, such as technological platforms for telerehabilitation. Use of these technologies was mostly said to represent a significant challenge for rehabilitation professionals to realize efficient care delivery, especially when there was a lack of technical support. This complexity was sometimes enough to discourage professionals from endorsing the innovation. One informant stated:

> *“We faced numerous IT-related limitations, just enormous. And you know, the support just isn’t there. (…) But I’ve completely let go of that—now I just manage with what we have and try to make things work.”* (Key informant 11)

Furthermore, the implementation of large-scale innovations requiring substantial changes and the involvement of numerous stakeholders was consistently associated with greater complexity. Most informants highlighted the specific challenges that emerged following the 2015 reorganization of the provincial healthcare system, which grouped together multiple territories and healthcare facilities (25,26). They explained how this restructuring amplified the complexity of communication channels, as well as administrative and decision-making processes:

> “*In 2015 […], several previously autonomous institutions came together. At that time, access mechanisms, service continuity processes, intake systems, and clinical practices were all different. […] So you understand that there’s a whole, whole, whole lot of complexity tied to that, beyond the clinical side*” (Key informant 7)

#### Barrier 3 - Insufficient resources to achieve successful implementation

Insufficient resources for implementing innovations was a recurrent barrier. This shortage was noticed at different stages in the implementation process, including planning and execution, which created an unfavorable environment for introducing new ideas. As expressed by this informant:

> *“We struggle to carry out clinical activities as they currently stand*. So, when we come and add a change, it’s difficult*. You know, adherence is difficult. But also we don’t have the possibility to free up someone, a clinical expert, to develop a [care] trajectory”* (Key informant 11).

In many organizations, the lack of qualified human resources needed to lead and provide the service was reported to have the most impact on implementation of the innovations. Moreover, during the COVID-19 pandemic, the organizations faced new challenges that shifted their priorities. This unprecedented situation generated a realignment of the innovations’ objectives. As stated by this informant:

> *“(…) the pandemic led to the reallocation of resources [to other services], practically putting an axe through the project itself because we had to redeploy resources. So the remaining resources had to be redirected to postoperative care, etc..”* (Key informant 3)

These challenges brought by the pandemic had long-lasting consequences on the potential for implementing innovations, related to persistent limited financial and human resources.

### Facilitator 1 - Matching evidence, contextual needs and resources

From the informants’ perspective, it was deemed essential to collect substantial evidence demonstrating that the innovation would yield the intended outcomes that were aligned with local needs. Such demonstration was instrumental in engaging rehabilitation professionals and enhancing their confidence in the change process. Reliable information sources included evidence-based literature and analogous strategies implemented in other clinical environments. One informant mentioned:

> *“Well, actually, we have literature that tells us how to proceed (…) We relied on the literature to be able to support the safety and relevance of our service offer. From there, we’ve implemented our own model.” (Key informant 9)*

In addition, involving qualified experts, such as researchers and internally appointed project developers, was reported as an efficient and reliable way to guide the planification and execution of the implementation process.

Proposing innovations that were adapted to each specific context, as well as population and professionals’ needs, was an important facilitator. One informant mentioned:

> *“I think that the fact of having prepared deployments by site, that took into account the reality of each of the sites, I think that’s one of the strong points. (…) I think it was really profitable to say to each of the sites: ‘(…) we will implement according to the reality of your site. What do you want? From what we’re offering? What works for you and what doesn’t?”* (Key informant 13)

### Facilitator 2 – Quality of the team involved in leading and conducting the implementation process

Many features related to the teams of individuals involved in the implementation processes were identified as important facilitators. The physical proximity of team members was said to facilitate the implementation and provide better support in the operations. No matter how many individuals were involved, relying on one appointed manager was considered as a key-point to facilitate decision-making process, and to successfully lead the organization towards implementation of the innovations. One informant stated:

> *“[Someone] who has the ability to think through the deployment steps and to create for himself an action plan (…), I think that’s essential.”* (Key informant 13)

The successful implementation of innovations was also significantly influenced by the intrinsic motivations and commitment of the professionals involved. When the innovations aligned with the core values of the practitioners, the process of implementation was considerably facilitated. Furthermore, effective implementation depended on the individual characteristics of team members, notably their expertise and clinical experience. As described by this informant:

> *“ Having served as a clinician and having experienced the realities of clinical practice also, allowed me to arrive not only with a macro-level perspective, but also having experienced it personally. So I had a good idea of what works, what doesn’t. That was really helpful.”* (Key informant 15)

## DISCUSSION

Through this environmental scan, we were able to identify and characterize various innovations that were implemented to improve access to outpatient rehabilitation for persons with physical disabilities, as well as to identify key barriers and facilitators to their implementation. These findings are important as they could be used to guide and optimize the development and implementation of future innovations in other rehabilitation settings that face access-related problems.

Using Levesque et al.’s framework (7) to describe underlying access issues, we identified innovations that aimed to improve a variety of services’ aspects. Waiting times before receiving outpatient rehabilitation services has received some attention in recent years (1,4,19,32), as it has detrimental effects on population health (33). Interestingly, the innovations identified through this environmental scan did not focus solely on excessive waiting times. Several innovations targeted other dimensions of access, such as geographic disparities or environmental limitations in accessing care during the COVID-19 pandemic—issues that also relate to service availability and accommodation (7). Other innovations aimed to improve the effectiveness of services (i.e., appropriateness according to Lévesque et al. 2011) or reduce inequities among specific subpopulations (i.e. acceptability) (7).

Among the identified innovations, different types of redesign strategies were operated, including the modification of care models, extending roles of rehabilitation professionals, or restructuring access processes. Most innovations resulted in small-scale, service-level changes, and only a few innovations were designed and implemented on a broader scale to optimize existing processes and improve overall system efficiency. These findings are consistent with other studies, highlighting the need for developing more systemic and integrated approaches to effectively address persistent access challenges (34,35).

In another vein, most innovations were implemented in primary care settings. This suggests that increasing access in such settings has received growing attention in recent years. Limited access to primary healthcare is indeed one of the main issues affecting health systems in many countries, and particularly in the province of Quebec (36). Innovations in primary care that we identified focused on rehabilitation professionals taking new roles, such as screening, triage and first-contact interventions (extending or modifying roles according to Dupuis et al.-(19)). These mostly aimed to address the negative effects of excessive waiting times before medical consultation, while helping to better meet the specific needs of adults with physical disabilities, such as those presenting musculoskeletal disorders. However, continued requirement to obtain a referral from a physician to consult a rehabilitation professional in many contexts limits timely access and undermines efforts to streamline care (37). Growing international evidence supports direct access to rehabilitation services and advanced practice models of care as an effective strategy to reduce wait times and disability, improve quality of life, and deliver cost-efficient care, by offering recovery-and prevention-oriented care (e.g., education and advice), often unavailable during standard medical consultations (38–41). In addition, while direct access to rehabilitation professionals is widely implemented in private settings in the province of Quebec, it is not usual practice in many publicly funded settings, although professional regulation does not require it.

Changes in the models of care delivery also represented a large proportion of the innovations implemented to improve access. Among these, new models of care increasingly incorporated e-health and digital innovations, a trend that was significantly accelerated during the COVID-19 pandemic (42). Restrictions regarding in-person care to minimize viral transmission prompted rapid adoption of technology-based solutions. While existing research supports the utilization of e-health in improving access to rehabilitation (43,44), our study highlights some barriers to implementation. The integration of technology-driven care models often introduced operational challenges for rehabilitation professionals, including increased administrative load and insufficient technical support, which at times led to disengagement from the innovation process. These findings suggest that, while e-health holds significant promise for addressing access barriers, its long-term success and sustainability will require more integrated, user-centered approaches that better account for the realities of clinical practice. Beyond digital health, additional strategies that modify care delivery—such as group-based interventions, joint interdisciplinary assessments, and providing more intensive interventions delivered over shorter periods have demonstrated potential to reduce waiting times and improve access to services (19). There may be room to further expand such strategies in Quebec.

### Strength and limitations

The strength of this project is the triangulation of different data sources and approaches. For example, the semi-structured interviews provided relevant additional content regarding the innovations and a deeper understanding about the nature of innovations, as well as the encountered facilitators and barriers. This study also has limits. To start, results are limited to innovations implemented in the province of Quebec, Canada, and may not reflect contexts elsewhere. It is also likely that, despite our efforts, our list of identified innovations may not be completely exhaustive. This may reflect the fact that healthcare-based innovations within public institutions are not systematically documented or do not explicitly state improving access as their primary goal (e.g. also aiming for better efficiency), which may have limited their identification. In addition, some of the targeted key informants were impossible to reach during the period in which the interviews were conducted. Hence, potential additional information related to the innovations could not be obtained. Furthermore, most of the identified innovations involved interdisciplinary or multidisciplinary care teams or physiotherapy professionals. Our findings, including identified barriers and facilitators, might not fully represent those encountered in other types of rehabilitation services. Also, because we were able to interview informants with different roles and backgrounds regarding the innovations, this most likely provided richness and diversity in the viewpoints but may have also introduced biases and prevented us from deepening certain aspects that a more homogeneous group would have shed light on.

## CONCLUSION

The results of this study advance our understanding of the nature and the specific purpose of innovations implemented to improve access to outpatient rehabilitation services for people with physical disabilities. Innovations targeting multiple barriers to access were identified, such as long waiting times, geographical inequalities and inappropriate services. The results of this study also highlight that the implementation of such innovations is complex and requires careful consideration of multiple factors. Further formal evaluation of innovations is also needed to help improve access to rehabilitation services.

## Data Availability

All data produced in the present study are available upon reasonable request to the authors

## Acknowledgements

The authors would like to thank all participating organizations and key informants who contributed their time and insights to this study. This work was supported by the following organizations through their respective grant programs for structuring projects: the *Centre interdisiciplinaire de recherche en réadaptation et intégration sociale (Cirris)*, as well as the *Alliance Santé Québec*.

