## Appendix 1 for "Innovations to improve access to outpatient rehabilitation services for persons presenting physical disabilities: results of an environmental scan in Quebec, Canada"

**Appendix 1:** Keywords guiding the online search strategy

| **Service** | **Accessibility** | **Innovation** |
| --- | --- | --- |
| Rehabilitation  Physiotherapy  Occupational therapy  Speech therapy  Psychology  « Social work »  Nutrition | Accessibility  Waiting  Access  Delay  Prioritization  List | Innovation  Improvement  Change |
